# Interleukin-33 stimulates stress in the endoplasmic reticulum of the human myometrium via an influx of calcium during initiation of labor

**DOI:** 10.1101/2021.11.05.21265965

**Authors:** Li Chen, Zhenzhen Song, Xiaowan Cao, Mingsong Fan, Yan Zhou, Guoying Zhang

## Abstract

**Background:** Inflammation is currently recognized as one of the major causes of premature delivery. As a member of the IL-1β family, interleukin-33 (IL-33) has been shown to be involved in a variety of pregnancy-related diseases. This study aims to investigate the potential function of IL-33 in uterine smooth muscle cells during labor.

**Methods:** Samples of myometrium from term pregnant (≥37 weeks gestation) women were frozen or cells were isolated and cultured. Immunohistochemistry and western blotting were used to identify the distribution of IL-33. Cultured cells were incubated with LPS to mimic inflammation as well as 4μ8C to block endoplasmic reticulum (ER) stress and BAPTA-AM, a calcium chelator.

**Results:** Similar with onset of labor, LPS could reduce the expression of nuclear IL-33 in a time-limited manner and induced ER stress. Meanwhile, Knockdown of IL-33 increased LPS-induced calcium concentration, ER stress and phosphorylation of NF-κB and p38. In addition, siRNA IL-33 further simulates LPS enhanced COX-2 expression via NF-κB and p38 pathways. IL-33 expression was decreased in the nucleus with the onset of labor. LPS induced ER stress and increased expression of the labor-associated gene, COX-2, as well as IL-6 and IL-8 in cultured myometrial cells. IL-33 also increased COX-2 expression. However, after IL-33 was knockdown, the stimulating effect of LPS on calcium was enhanced. 4μ8C also inhibited the expression of COX-2 markedly. The expression of calcium channels on the membrane and intracellular free calcium ion were both increased accompanied by phosphorylated NF-κB and p38.

**Conclusions:** These data suggest that IL-33 may be involved in initiation of labor by leading to stress of the ER via an influx of calcium ions in human uterine smooth muscle cells.

**Funding:** This study was supported by grants from the National Natural Science Foundation of China (Nos. 81300507).

## Introduction

Preterm delivery occurs before 37 weeks of gestation and is one of the major causes of perinatal morbidity and mortality^[1]^. The main difference between preterm and term labor is when the initiation of labor begins. It is widely accepted that infection, uterine over-distension, decline in progesterone action, breakdown of maternal-fetal tolerance, stress and other unknown reasons are involved in preterm labor^[2]^. However, all of these reasons lead to inflammatory events. These are mainly due to the identification of microorganisms and their products, and increased secretion of pro-inflammatory cytokines and chemokines in the decidua and myometrium, which enable leukocytes to infiltrate the uterus leading to its activation ^[3, 4]^.

As a member of IL-1 family, IL-1β is one of the well-recognized inflammatory factors that induce the initiation of labor. Interleukin-33 (IL-33) is a member of the IL-1β family and is expressed in a variety of cells, including innate immune (macrophage and dendritic), Th2, B, epithelial, endothelial and muscle cells. Also, it has been shown to have a key role in various diseases, including inflammatory, autoimmune (including asthma), cardiovascular and allergic diseases^[5, 6]^.

Recently, Huang et al. found that after exogenous administration of IL-33, it was able to activate B cell-mediated immune regulation and promote the expression of progesterone-induced blocking factor 1 (PIBF1), which plays an essential role in safeguarding against preterm labor^[7]^. Preliminary data showed that IL-33 was mainly located in the nuclei of myometrial cells, and with the onset of labor, IL-33 expression in the nucleus decreased. Studies also showed that mast cells responded to the secretion of IL-33 after exposure to the allergens and this affected airway smooth muscle hyper-responsiveness by promoting the contraction and enhancing calcium influx of airway smooth muscle, leading asthma attacks^[8, 9]^. In addition, IL-33 was found to be effective in alleviating the myocardial endoplasmic reticulum (ER) stress response caused by myocardial injury^[10, 11]^. These studies suggested a potential role of IL-33 in human parturition and prompted us to investigate the changes in IL-33 expression in the nucleus of the myometrium during the third trimester of pregnancy.

## Results

### Nuclear expression of IL-33 is reduced during labor

In order to establish the nuclear function of IL-33, we first confirmed the presence of the nuclear protein in human uterine smooth muscle cells. Immunohistochemistry with the anti-IL-33 antibody revealed that IL-33 was expressed in the nuclei of myometrial cells. The term labor (TL) and preterm labor (PTL) groups showed weak staining of IL-33 in the nuclei of myometrium cells. In contrast, strong nuclear staining of IL-33 was observed in the myometrial cells of term nonlabor (TNL) and preterm nonlabor (PNL) groups (Figure 1A).

**Figure 1.**
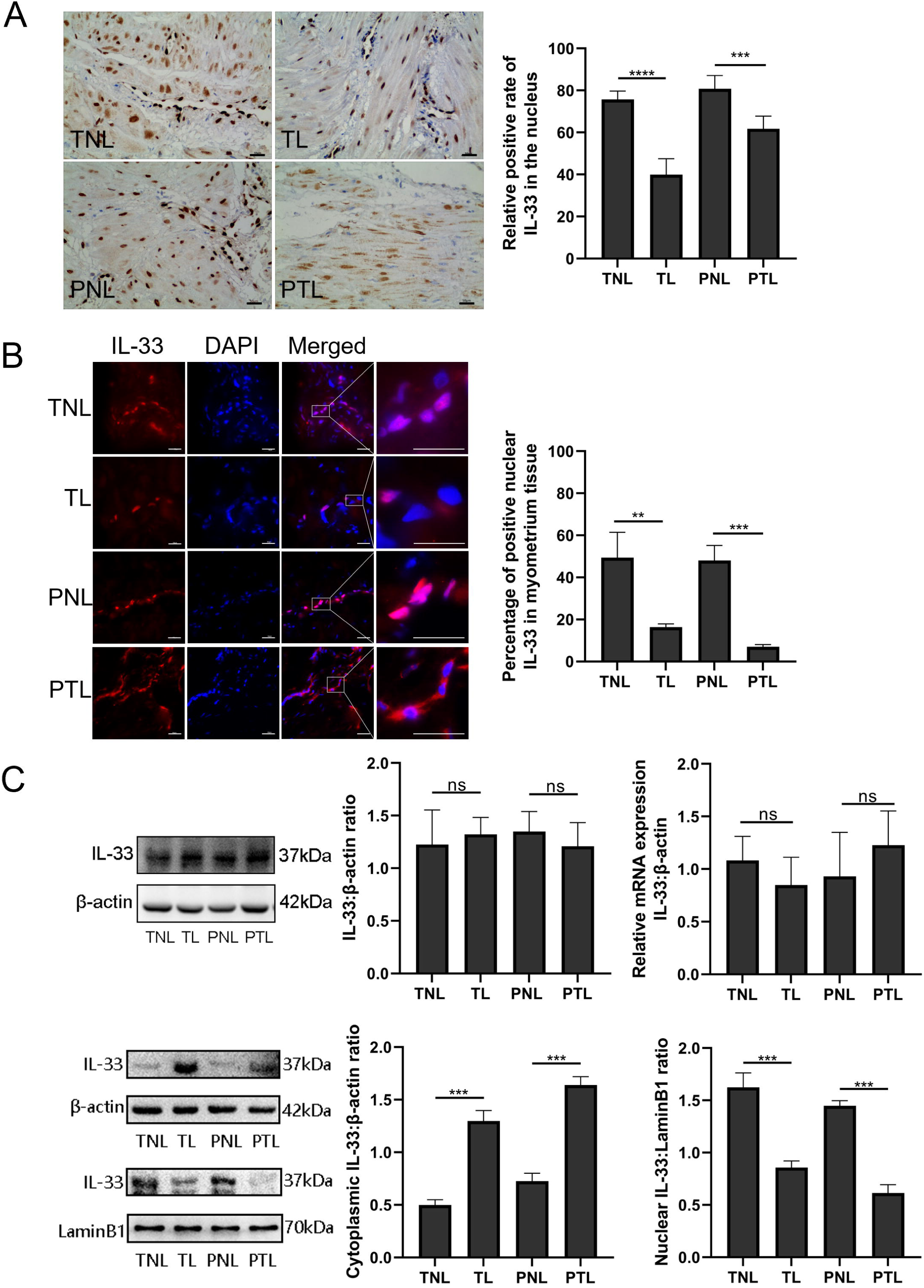
Nuclear localization of the IL-33 and reduced with the onset of labor. The sections fixed paraffin-embedded of human myometrium were immune-stained with anti-IL-33 antibody. (A) Immunohistochemistry results showed that whether in TNL or PNL tissue, IL-33 mostly located in the nuclear while reduced sharply and emerged in the cytoplasm with the initiation of labor as shown in figure TL and PTL. Quantification of IL-33 expression within the nucleus region showed apparent differences between labor and non-labor tissue (n=4). (B) The sections of human myometrium were visualized by an Alexa Flour 594 secondary antibody labeled with IL-33 (Red), and nuclei were stained with DAPI (Blue). Analysis of nuclei was performed by confocal microscopy, fluorescence signals of IL-33 and nuclei were superimposed. Immunofluorescence staining results reflected the same phenomenon. (C) From Western blots, we also can see that non-labor groups had more nuclear expression and less cytoplasmic expression of IL-33 compared to labor groups while the total IL-33 had no obvious differences between groups. QT-PCR discovered no apparent alteration in the levels of IL-33 mRNA. Each value represents the mean ± standard deviation (SD) of three independent experiments. ** P<0.01, *** P<0.001, bar 50 μm.

In order to identify the localization of IL-33 changes during labor, immunofluorescence staining was performed on myometrium sections. This also confirmed the nuclear localization of IL-33 in the myometrium cells. It should be noted that the expression of nuclear IL-33 was more intense in the TNL and PNL groups when compared with TL and PTL groups. There were similar levels of staining in the two no labor groups as well as the two labor groups (Figure 1B). Western blotting analysis revealed that there was no difference in the expression of total IL-33 among all groups. To confirm the nuclear localization of the IL-33, we separated the cytoplasmic and nuclear fraction of myometrial cells. There was decreased levels of nuclear IL-33 from the TNL to the TL groups, which was consistent with the change seem from PNL to PTL. The results of qPCR suggested that mRNA expression of total IL-33 was not different between the different groups (Figure 1C).

### Treatment with LPS resulted in a time-dependent nuclear IL-33 decrease

To examine the function and the mechanisms of nuclear IL-33 during parturition, myometrial cells were incubated with LPS and the kinetic changes of IL-33 we visualized by confocal microscopy. Following treatment with LPS, there was a marked decrease in endonuclear IL-33 levels for approximately 3 hours and then increased to above control values by 12 hours, which was higher than that in the control group (Figure 2A). For the following 12h the levels appeared to decrease again. We next investigated the effect of LPS on the localization of IL-33 in these cells. Western blots confirmed that the expression levels of total IL-33 were reduced within 6 hours, then peaked at 12 hours. The cytoplasmic and nuclear fractions of primary myometrial cells were also isolated at different time points after LPS treatment. Western blot analysis showed that nuclear IL-33 reduced with the stimulation by LPS. However, the levels again peaked in the nucleus at 12 hours, and was higher than seen in control cells (Figure 2B).The above shows that LPS induces a time-dependent reduction of IL-33 in the nucleus.

**Figure 2.**
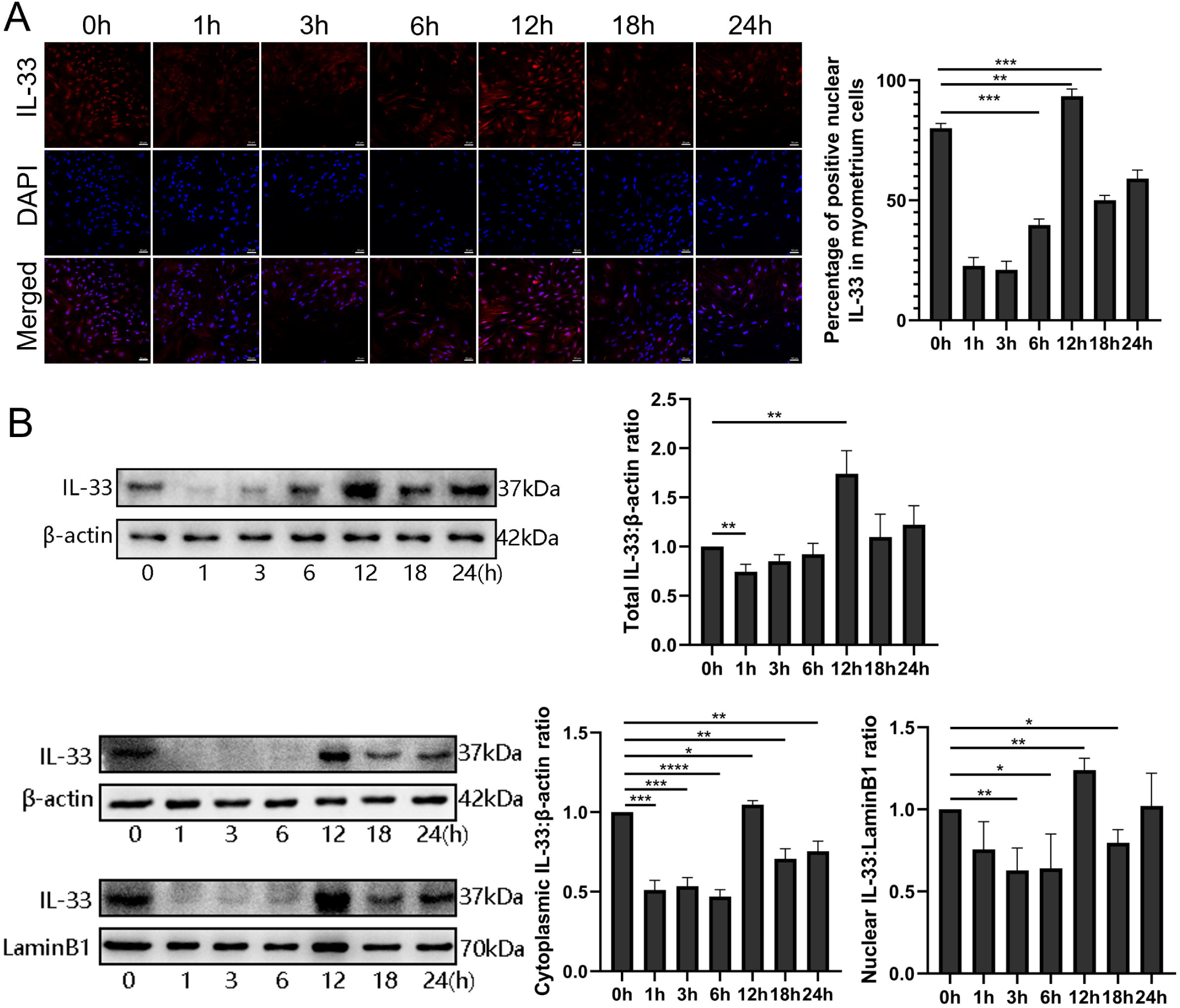
Dynamic changes of IL-33 position during LPS stimulation. (A) Localization analysis of confocal laser scanning microscopy images of IL-33 with the stimulation of 10 μg/ml LPS, we could see that IL-33 in the nucleus declined sharply compared with the control group, especially in 3 hours. However, with the longer time of the LPS treatment, the expression of IL-33 commenced to rise again from 6 hours and reached the peak at 12 hours. The lower panel is the percentage of cells of which IL-33 expressed mainly in the nucleus (n=3). (B) After the primary cells were loaded with LPS for different times, cytoplasmic and nuclear proteins were isolated. In the cytoplasmic and nucleus fraction IL-33 were quantified by Western blotting, the experiment was performed as described for Figure 2B. Each experiment was performed at least three times while shown are representative results (n=3). From the blots, we known that whether cytoplasmic or nucleus fraction IL-33 levels presented the same change trend as immunofluorescence. * P<0.05, ** P<0.01, bar 50 μm.

### Silencing of IL-33 enhances LPS-induced calcium ion levels

To investigate whether the presence of IL-33 regulates uterine smooth muscle contraction, knockdown experiments targeting IL-33 were performed before LPS treatment for 6 hours or 12 hours, respectively. After stimulating for 6 hours, the increase in intracellular calcium levels was observed by using a laser confocal microscope to observe intracellular fluorescent probes which was consistent with the calcium channels status (Figure 3A). Apparent enhancement in protein levels of calcium channels proteins, Cav3.1 and Cav3.2, were demonstrated after siRNA-based LPS treatment while siRNA-mediated efficient knockdown of IL-33 was simultaneously observed (Figure 3B).

**Figure 3.**
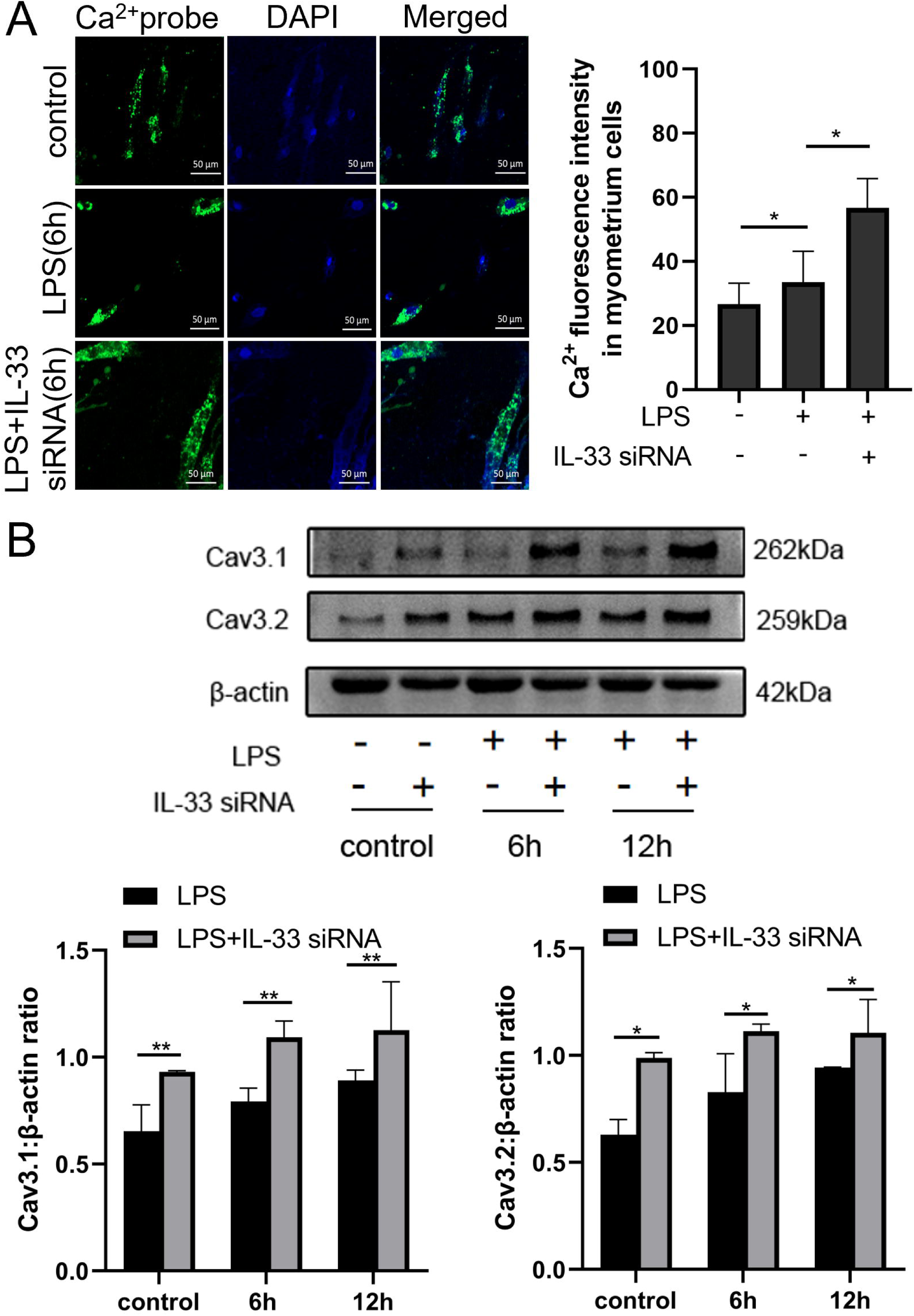
IL-33 silencing enhanced LPS-induced expression of calcium channels and the intracellular calcium concentration. Primary myometrium cells were transfected with IL-33 or non-targeting (as control) siRNAs and treated with 10 μg/ml LPS or PBS (as control) for 6 hours. (A) Immunofluorescence staining analysis performed with fluo-3AM (green), nuclei were stained with DAPI (blue) (n=3). (B) Western blot analysis for expression levels of Cav 3.1 and Cav 3.2 in stably transfected and non-transfected myometrium cells with treatment as indicated (n=5). * P<0.05, ** P <0.01, bar 50 μm.

### Inhibition of cytoplasmic calcium attenuates LPS induced ER and COX-2

LPS was able to induce cytoplasmic calcium levels and ER stress in uterine smooth muscle cells, but whether there is a correlation between the two is unknown. In this study, BAPTA-AM was used to evaluate the role of elevated cytoplasmic calcium levels during ER stress. The results showed an attenuation in the expression of LPS-induced COX-2 after 12 hours of preloading cells with BAPTA-AM (Figure 4A). After LPS stimulation for 6 and 12 hours respectively, the intracellular ER state was assessed by western blotting analysis. We found that p-IRE1α and XBP1s in the myocytes of pre-chelating-based stimulation showed a decreasing trend when compared with cells stimulated by LPS directly. Furthermore, on the basis of that no significant difference was detected in LPS treatment when directly compared to controls, there was no apparent alteration in protein levels of GRP78 in pre-chelating-based stimulation (Figure 4B).

**Figure 4.**
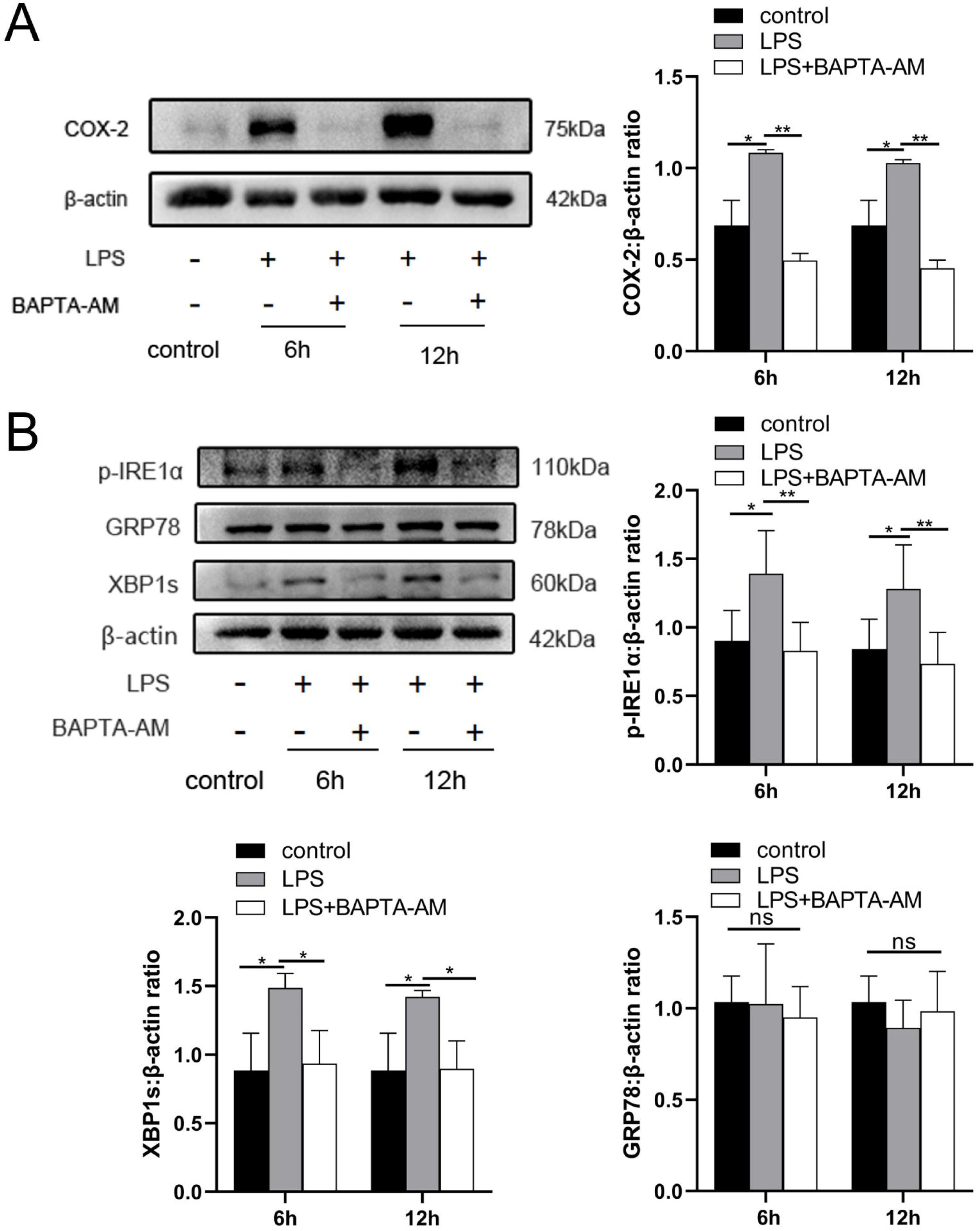
BAPTA-AM inhibited expression of COX-2 and ER stress response. (A) Study of COX-2 protein expression with LPS treatment in the absence or presence of BAPTA-AM (25μg/ml) (n=5). (B) The protein expression of p-IRE1α, XBP1s and GRP78 were analyzed by Western blot at 6 hours and 12 hours under LPS stimulate and with or without BAPTA-AM (n=5). * P<0.05, ** P<0.01.

### IL-33 affects the LPS-induced ER stress response in myometrium cells

Since studies have indicated a crucial role of ER stress during successful pregnancies as well as in the pathogenesis of preeclampsia, we decided to assess whether the ER stress response also played a role in labor. As shown in Figure 5A, protein expression of the ER stress□sensing molecules, p-IRE1α and XBP1s, were significantly increased in the TL and PTL groups when compared with the TNL and PNL groups. However, there was no significant change in GRP78 protein expression among the groups.

**Figure 5.**
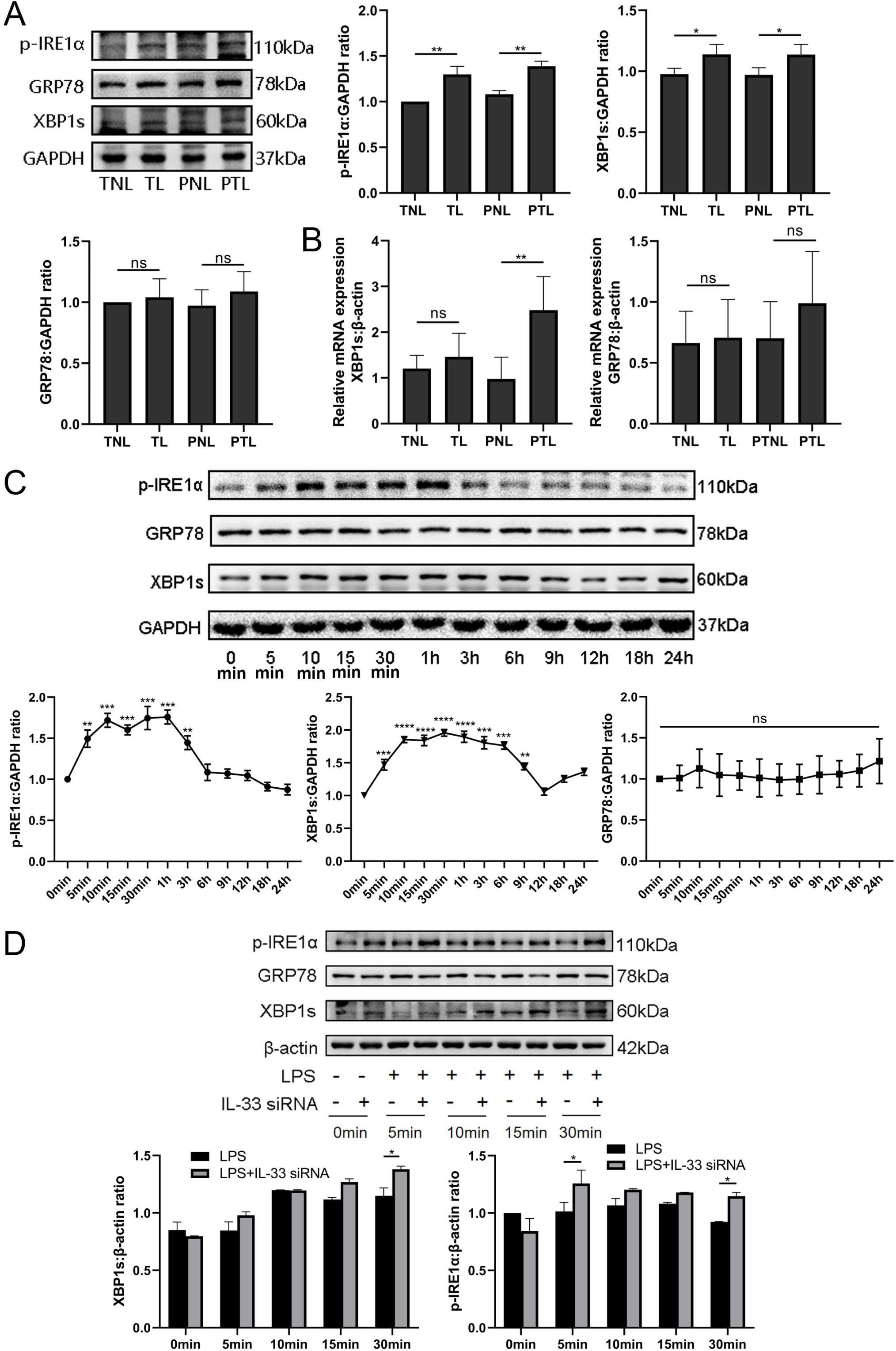
IL-33 silencing highlighted LPS-induced endoplasmic reticulum stress response. Based on the discovery that calcium ions affected endoplasmic reticulum stress, Western blot and QT-PCR were we further to explore the expression of endoplasmic reticulum stress in tissues from protein and mRNA level. (A) The protein levels of P-IRE1α and XBP1s in the TL and PTL groups were higher than those in the TNL and PNL groups while there was no alteration in the level of GRP78 protein(n=6). (B) The mRNA level of XBP1s in the PTL groups was higher than that in the PNL groups (n=6). Furthermore, in order to illustrate the protein level changes of endoplasmic reticulum stress during labor, LPS was used to stimulate primary uterine smooth muscle cells for different time course and then Western blot was used to detect the alteration of endoplasmic reticulum stress. (C) It was found that the protein level of pIRE1α reached its peak at 10 minutes while XBP1s at 15 minutes, while the protein level of GRP78 did not change significantly (n=5). We also detected apparent alteration in endoplasmic reticulum stress protein during cells stimulated based knockdown experiments targeting IL-33. (D) It revealed that the endoplasmic reticulum stress response in the siRNA-based group was more obvious compared with the LPS stimulated directly especially at 30 minutes (n=5). * P<0.05, ** P<0.01, *** P<0.005, ****P<0.001

In order to verify whether the transcription levels changed, qPCR results were compared those of western blotting analysis and were found to be similar (Figures 5A and 5B). Q-PCR showed that the mRNA expression of GRP78 had no intergroup difference. However, XBP1s mRNA expression was increased in PTL when compared with PNL, but there was no difference between the TNL and TL groups. Overall, these findings support the hypothesis that activation of ER stress is a potential mechanism underlying labor. In order to confirm this, we stimulated primary myometrial cells with LPS and induced the ER stress response without altering GRP78 protein levels. We found that the expression levels of p-IRE1α and XBP1s reached their peaks at 1 hour and 30 minutes, then decreased to almost the basal levels at 6 hours and 12 hours, respectively (Figure 5C). We next suppressed its expression with IL-33 siRNA in these cells and found that IL-33 knockdown heightened the LPS-induced ER stress response. This is shown in Figure 5D and in contrast to the LPS group, XBP1s and phosphorylated IRE1αprotein levels were increased in the IL-33 siRNA group.

### IL-33 siRNA and ER stress response regulate COX-2 expression in myometrium cells

In order to study whether IL-33 has a direct impact on expression of COX-2, we introduced IL-33 siRNA into primary myometrial cells and treated them with LPS for 12 hours. COX-2 was up-regulated of in the IL-33 siRNA group (Figure 6A). The previous results showed that a milder ER stress response occurred with the onset of labor and could be modulated by cytoplasmic calcium levels. To define the possible effect of the ER stress during the labor, the IRE1 inhibitor, 4μ8c, was used to block the LPS-induced ER stress response. Western blots were conducted to examine whether the expression of COX-2 could be inhibited. The result demonstrated that 4μ8C inhibited the expression of COX-2 markedly (Figure 6B).

**Figure 6.**
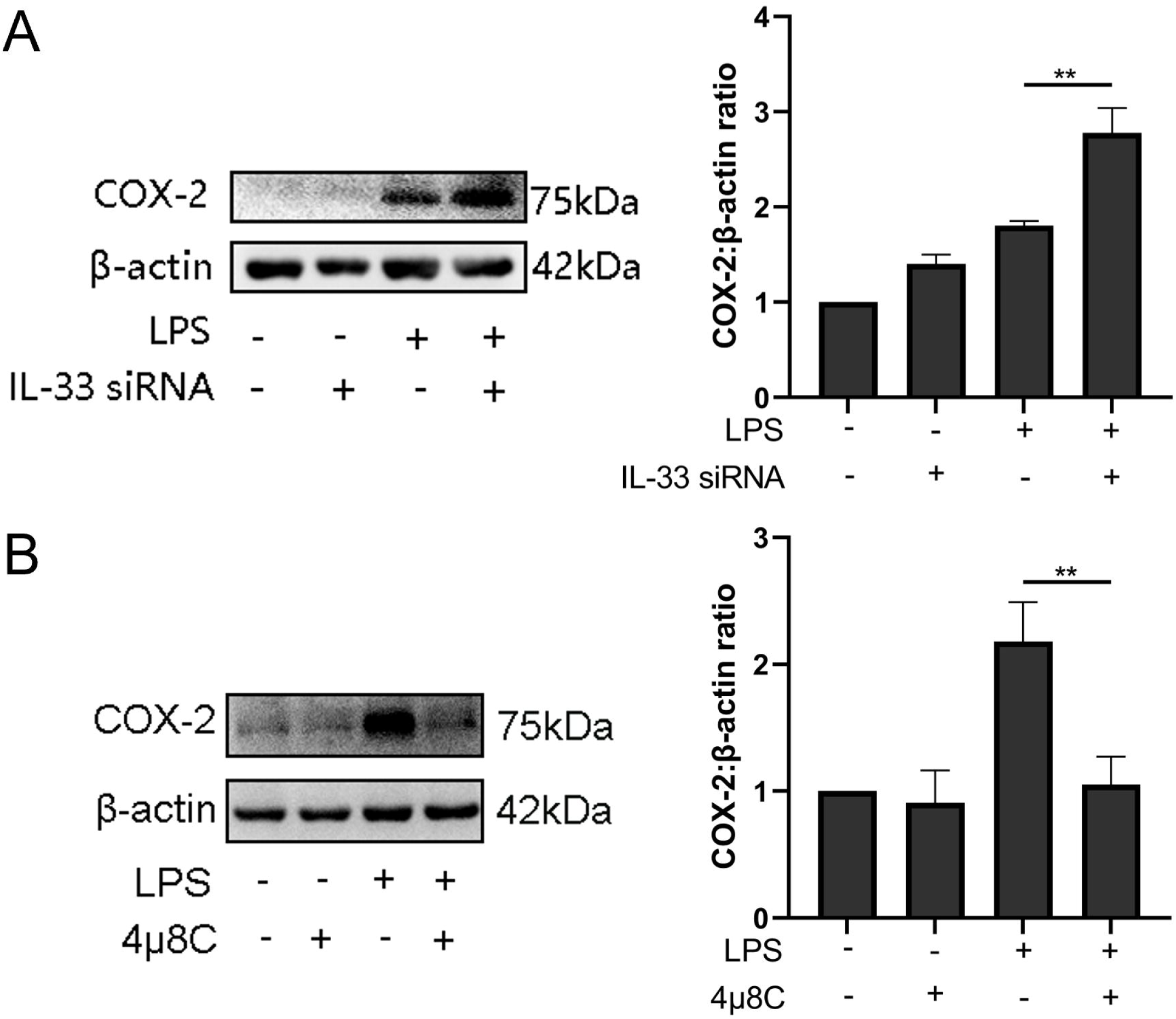
IL-33 siRNA and ER stress response affected COX-2 expression in myometrium cells. (A) In the process of studying whether IL-33 affects COX-2, we found that the COX-2 expression in the siRNA-mediated group was significantly increased compared with the LPS alone group (n=5). (B) Western blot analyses showing protein expression of COX-2 in myometrium cells was decreased following treatment with LPS for 12 hours (n=5). * P<0.05, ** P<0.01. Each value represents the mean ± standard deviation (SD) of three independent experiments.

### IL-33 affects COX-2 expression via p38/MAPK and NF-kB signaling pathways

We also found the expression of phosphorylation of p38 and NF-κB increased significantly when LPS caused an induction of COX-2 expression, and these peaked at 30 minutes (Figure 7A). Based on above observations, we wondered whether IL-33 was involved in these signaling pathways, thereby affecting the expression of COX-2. Based on the knockdown experiments targeting IL-33 which showed that when siRNA-based LPS stimulation lasted for 1 hour, the protein levels of NF-κB phosphorylation were significantly increased compared to that of the group without IL-33 knockout. However, the phosphorylation of p38 was not significantly different within 30 minutes of LPS stimulus but elevated sharply when the stimulus lasted for 30 minutes (Figure 7B). In addition, p38/MAPK and NF-KB signaling pathway inhibitors, SB-202190 and JSH-23 respectively, were added before LPS treatment, and western blotting analysis showed that COX-2 expression was significantly reduced (Figure 7C).

**Figure 7.**
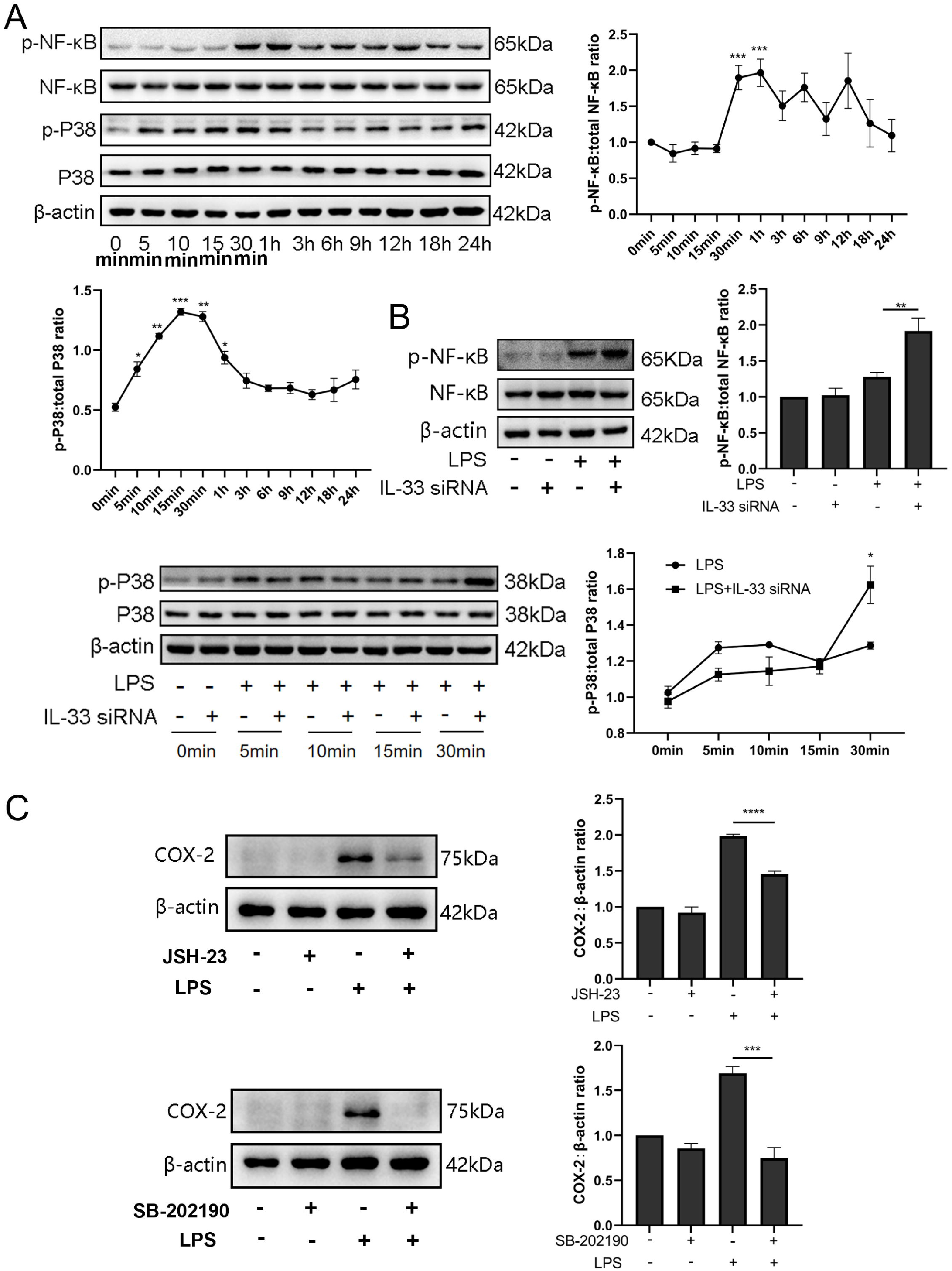
IL-33 knockdown enhanced LPS-induced NF-κB and p38/MAPK signaling pathways. (A) Relative levels of p-P38, P38, p-NF-κB and NF-κB were assessed by western blot analysis at the indicated time point after LPS (10 μg/ml) stimulation. Phosphorylation levels of P38 and NF-κB increased gradually with LPS stimulation and peaked at 15 minutes and 1 hour, respectively(n=5). (B) Western blot analysis of p-P38, P38, p-NF-κB and NF-κB expression in cells transfected with siRNA targeting IL-33 after treatment with LPS for 30minutes. Compared with the LPS group, the protein expression of phosphorylated P38 and NF-κB were increased in the LPS + siRNA IL-33 group(n=5). (C) The protein level of COX-2 was decreased when SB-202190 and JSH-23 blocked p38/MAPK and NF-KB signaling pathway, respectively(n=5). * P<0.05, ** P<0.01, *** P<0.001.

### IL-33 siRNA and cytoplasmic calcium influence the expression of IL-8 and IL-6 secreted by myometrium cells

The expression levels of IL-8 and IL-6 in the supernatants of the LPS treated cells were increased when compared with the controls. In addition, in comparison with the LPS group, more IL-8 and IL-6 were released into the cell supernatants after IL-33 siRNA transfection and partial binding of cytoplasmic calcium ions (Figure 8).

**Figure 8.**
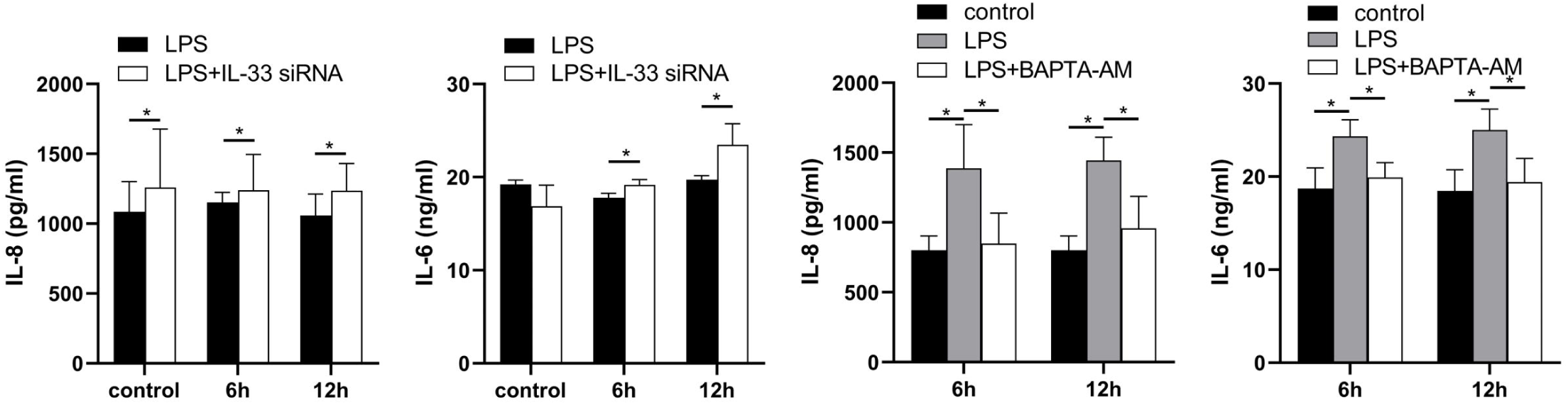
IL-33 siRNA and cytoplasmic calcium influence the expression of IL-8 and IL-6. Compared with the LPS group, the expression of IL-8 and IL-6 was increased in the LPS+IL-33 siRNA group and decreased in the LPS+BAPTA-AM group (n=5). * P <0.05.

## Discussion

Although the specific mechanism of premature delivery is still unclear, inflammation is currently recognized as one of the major causes of this phenomenon^[12, 13]^. The current understanding of this process is that as the myometrium switches from a quiescent to a contractile state, it also undergoes a shift in signaling from anti-inflammatory to pro-inflammatory pathways^[14, 15]^. As an inflammatory factor, IL-33 has many similar effects to IL-1β. IL-33 is composed of a N-terminal nuclear domain and a C-terminal IL-1-like cytokine domain which is mainly localized in the nucleus and binds to histones and chromatin^[16]^. It proposed that IL-33 plays a dual role, playing a pro-inflammatory role as an inflammatory cytokine outside the cell and participating in the regulation of transcription as a nuclear factor^[17]^.

Numerous studies have shown that IL-33 had an essential role in pregnancy-related diseases such as recurrent abortion and preeclampsia^[18, 19]^. In early pregnancy, IL-33 can increase the proliferation and invasion of the decidua, and decidual macrophage-derived IL-33 is a key factor for placental growth, and its deletion can lead to adverse pregnancy outcomes^[20, 21]^. Similarly, IL-33 expression is reduced in the placenta of preeclampsia patients, which affects placental function by reducing the proliferation, migration and invasion of trophoblast cells^[22, 23]^. In preliminary experiments, we found that IL-33 was expressed in the nuclei of myometrium cells in the third trimester of pregnancy and the levels decrease following the onset of the labor. This was consistent with ta previous study which showed that IL-33, a mucosal alarmin, was being sequestered in the nucleus in order to limit its pro-inflammatory potential. It was released as a warning signal when tissue was damaged following stress or infection such as its release from the nucleus during asthma attack^[9, 24]^.

In order to identify factors that are important in driving PTL progression, we utilized LPS to stimulate primary cultures of uterine smooth muscle cells to mimick inflammation during labor. This resulted in a rapid decrease of IL-33 in the nucleus after LPS treatment which was ameliorated after the stimulus duration was lengthened. It is well known that labor is a physiological process and will not cause a maternal pathological state, so we hypothesized that the recruitment of IL-33 may be to maintain cell homeostasis and enable its normal function. In order to explore the potential effect of nuclear IL-33 during labor, we knocked down IL-33 in primary myometrial cells before LPS stimulation. Initially, we found that after introduction of siRNA IL-33, the intracellular calcium ion levels and the expression of T-type calcium channel proteins (Cav3.1 and Cav3.2) on the membrane increased sharply. This was consistent with other studies which showed that IL-33 can affect calcium elevation autonomously or it can cooperate with other mediators to achieve this effect, although the specific mechanism is still unclear^[25]^.

Myometrial contractions are mainly regulated by intracellular Ca^2 +^ levels and calmodulin and the effect of IL-33 on calcium elevation is likely to be a direct effect upon myometrial tissues^[26]^. However, we found that reduced IL-33 in the nucleus promoted the release of pro-inflammatory cytokines such as IL-8 and IL-6 in myometrial cells, while the expression of pro-inflammatory cytokines was reduced after Ca2^+^ binding in the cytoplasm. This led us to conclude that IL-33 does play a certain role in the process of parturition. Intracellular calcium was closely linked to the ER stress response. It is well documented that when tissues and cells are irretrievably stimulated or damaged, the ER stress response occurs to maintain cell homeostasis. The ER which is the main intracellular calcium storage chamber, plays a vital impact on maintaining Ca2+ homeostasis in various organelles, and homeostasis of the ER is closely related to intracellular Ca2+ concentration^[27]^.

Researchers recently found that LPS stimulation of uterine explants could cause an ER stress response in uterine muscle. This led them to hypothesized that dysregulation of ER homeostasis in uterine smooth muscle might be one of the mechanisms of LPS induced inflammatory preterm delivery^[28]^. When compared with non-laboring specimens, the levels of GRP78, IRE1 and XBP1s in fetal membranes and myometrium were elevated in both TL and PTL^[29]^. Consistently, the expression of the ER stress response in the uterine smooth muscle was increased during labor when compared to non-labor tissues. Here, we found that, although there was no obvious change in GRP78, knock down of IL-33 increased the expression of IRE1α and XBP1s significantly. As a chaperone protein in the ER, GRP78 binds to three prominent ER stress proteins under steady state conditions. These are PERK, ATF6 related to redox reaction and the Golgi apparatus, respectively. An additional protein, IRE1α, is also involved and it is related to inflammatory signals and repairing protein mis-folding^[30, 31]^. During the adaptive phase of the ER stress response, the IRE1α-XBP1s pathway is activated.

To explore the potential mechanism of how IL-33 is involved in the process of labor, we transfected siRNA IL-33 into primary myometrial cells, and the decrease in nuclear IL-33 prompted the increase of COX-2 expression after LPS stimulation. However, after IL-33 was knocked out, the stimulating effect of LPS on COX-2 which could be blocked by calcium chelating agents was more obvious. Furthermore, we explored whether the modest ER stress response seen relates to the process of labor. We found that the expression of COX-2 was sharply decreased by loading the primary cells with an ER stress response blocker which is similar with research results seen with modulation of pain by leukocytes^[32]^.

Based on the above findings, we speculated that IL-33 could influence COX-2 expression via two pathways: 1) it directly prevented excessive COX-2 expression in myometrial cells after LPS stimulation and 2) it influenced COX-2 expression by maintaining the severity of ER stress response. The binding of IL-33 to NF-κB in the nucleus could block NF-κB signaling and inhibit the transduction of downstream associated inflammatory signals^[33]^. Consistent with previous studies^[34]^, phosphorylation of NF-κB by LPS treatment was increased after IL-33 knockdown in myometrial cells. Also, we found that a change of the p38/MAPK signaling pathway was consistent with that of the NF-κB signaling pathway. In addition, selective blocking of NF-κB and p38/MAPK signaling pathways significantly inhibited the expression of COX-2. Therefore, we hypothesized that NF-κB and p38/MAPK signaling pathways may be the potential downstream pathways of IL-33 affecting LPS-induced COX-2 production (Figure 9).

**Figure 9.**
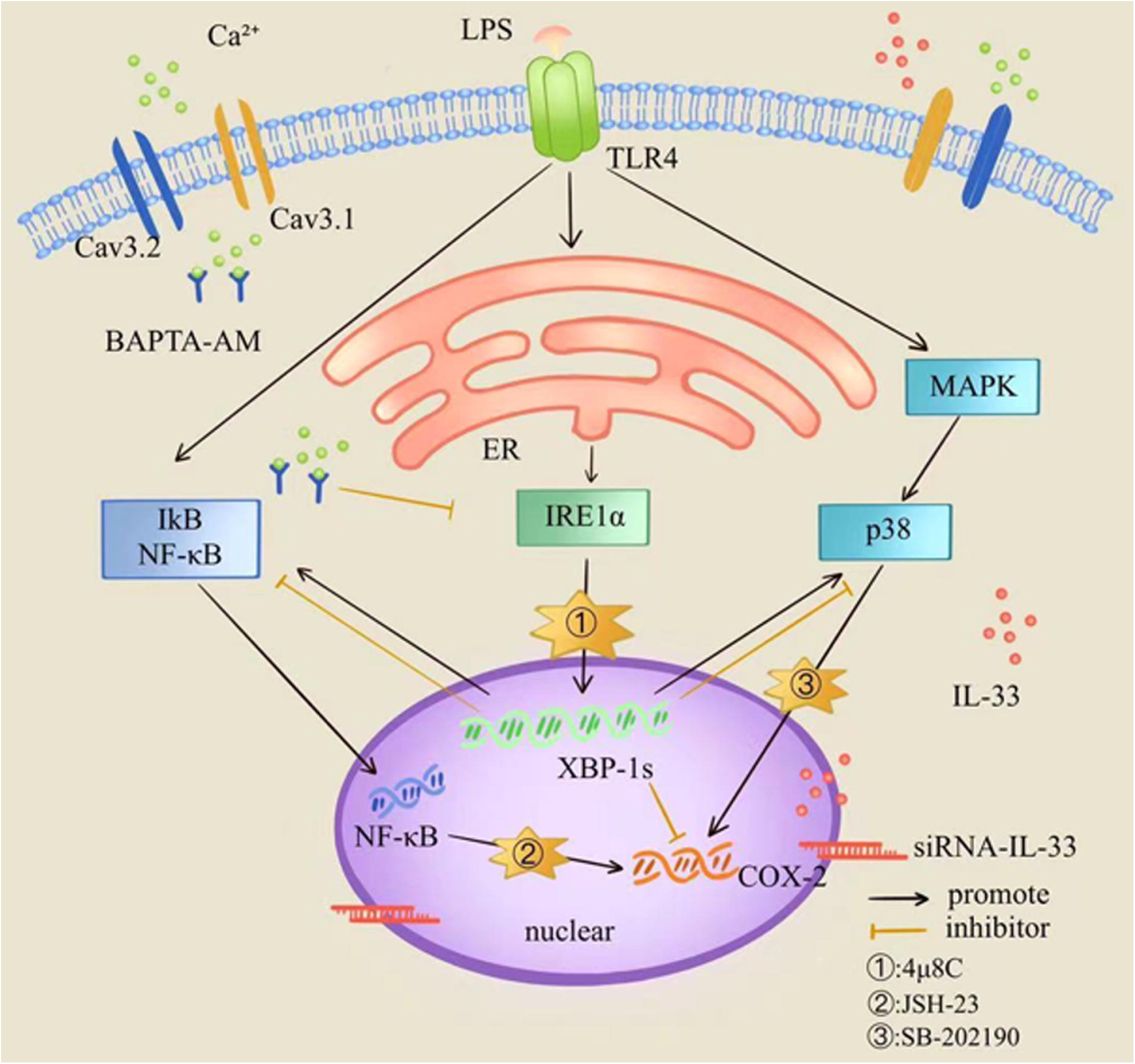
Model for the role of IL-33 in the myometrium participates in maintaining a uterine quiescent state at the tissue-to-cellular level during late pregnancy.

To summarize, we found that the nuclear factor, IL-33, can affect contraction of uterine muscle cells so as to participate in parturition by regulating intracellular calcium ion and thus influencing the ER stress. Our study demonstrates that nuclear IL-33 in the myometrium participates in maintaining a uterine quiescent state at the tissue-to-cellular level during late pregnancy. Further studies on whether IL-33 is involved in the redox reaction of mitochondria as well as regulation of intracellular calcium ions caused by ER stress are necessary.

## Materials and Methods

### Ethics

The subjects of the study were cesarean section patients in the Obstetrics Department of the First Affiliated Hospital of Nanjing Medical University. These included only cesarean section patients without obstetric complications, such as breeched pregnancies, mothers with scarred uteruses and giant babies. All patients signed an informed consent form. The study was approved by the Ethics Committee of the First Affiliated Hospital of Nanjing Medical University, China, and complies with the Declaration of Principles of Helsinki.

### Specimen collection

Patients were separated into 4 groups: term labor (TL), preterm labor (PTL), term non-labor (TNL) and preterm non-labor (PNL) groups. When undergoing cesarean section, a sample of the uterine smooth muscle (1.0×1.0×1.0cm^3^) was cut from the site of incision at the lower and upper edge of the uterus. The muscle was washed 3 times in pre-cooled PBS to remove excess blood, quickly frozen in liquid nitrogen, and stored at -80□. In some cases, similar sized samples were stored in cold PBS and used for culture of myometrial smooth muscle cells.

### Western blotting

Proteins were extracted from human primary myometrium cells and human uterine smooth muscle tissues. Proteins were separated by using 8% or 10% sodium dodecyl sulfate polyacrylamide gel electrophoresis and transferred to polyvinylidene fluoride membranes (IPVH00010, Mreck Millipore). Nonspecific binding sites were blocked by incubation with 5% non-fat milk for 2 hours. Membranes were then incubated with specific antibodies overnight at 4°C and washed 3 times with phosphate-buffered saline with Tween-20 (PBST) and then incubated with infrared dye–labeled secondary antibodies for 2 hours at room temperature.

After further washes with PBST, membranes were visualized using a Bio-Rad gel imager and the results were normalized to β-actin or GAPDH protein and expressed in arbitrary units. The antibodies used were: anti-IL-33 antibody (1:1000, ab54385, Abcam); anti-p-IRE1α antibody (1:1000, ab124945, Abcam);anti-XBP1s antibody (1:1000, 83418, Cell Signaling Technology); anti-GRP78 antibody (1:1000, 11587-1-AP, Proteintech); anti-COX-2 antibody (1:1000, ab179800, Abcam); anti-p38 antibody (1:1000, D13E1, 8690, Cell Signaling Technology); anti-NF-κB antibody (1:1000, D14E12, 8242, Cell Signaling Technology); anti-phospho-p38 antibody (1:1000, D3F9, 4511, Cell Signaling Technology); anti-phospho-NF-κB antibody (1:1000, 93H1, 3033, Cell Signaling Technology); anti-Cav 3.1 antibody (1:2000, DF10014, Affinity) and anti-Cav 3.2 antibody (1:500, sc-377510, Santa Cruz Biotechnology).

### Immunohistochemistry

The fixed paraffin-embedded sections of human myometrium were rehydrated in a graded series of decreasing alcohol concentrations. Sections were incubated in 3% hydrogen peroxide for 30 minutes to block endogenous peroxidase and 2% normal goat serum for 1 hour to reduce nonspecific binding. Then samples were incubated overnight at 4□ with goat anti-human IL-33 monoclonal antibody (MAB36253, R&D) at a concentration of 0.1 µg/mL and then 1-3 drops of biotinylated secondary antibodies for 2 hours at room temperature. 1-5 drops of DAB chromogen solution were added to cover the entire tissue section and incubated for 10 minutes. The sections were rinsed in deionized H2O and the slides were drained and the stained tissue was covered with a coverslip of an appropriate size. These were then visualized under a microscope using a bright-field illumination.

### Real-time quantitative PCR (qPCR)

Total RNA was isolated using Trizol reagent (SN114, NanJing SunShine Biotechnology). The RNA was reverse transcribed to cDNA using a cDNA first strand synthesis kit (D6110A, Takara Bio), according to the manufacturer’s instructions. The upstream and downstream primer sequences used are shown in Supplemental Table 1. Quantitative, real-time PCR was then conducted with SsfastTMEvaGreen Supermix (172-5201AP, BIO-RAD) in a BioRad CFX96 Touch Real-Time PCR. The relative abundance of mRNA in myometrium was normalized to that of β-actin using the comparative cycle threshold method (2−ΔΔCT).

### Immunofluorescence

Tissue sections and cells were fixed with 4% paraformaldehyde in PBS for 30 minutes at room temperature, washed three times with PBS and repaired with 75% glycine for 10 minutes. Then they permeabilized with 0.5% Triton X□100 in PBS for 30 minutes, and then washed three times with PBS and blocked with 3% bovine serum albumin (BSA, sh30087.02, HyClone) in PBS for 1 hour. These operations were conducted at room temperature. Primary antibody incubation was performed overnight at 4°C with goat monoclonal anti-IL-33 (1:200, MAB36253, R&D) diluted in 0.2% BSA in PBS. For detection, the tissues were incubated for 2 hours at room temperature with Alexa Flour 594 secondary antibody (1:500, A32758, Life Technologies) which was diluted in 0.2% BSA in PBS. The samples were washed as described above, mounted using Antifade Mounting Medium with DAPI (P0131, Beyotime Biotechnology) and analyzed using a confocal fluorescence microscope (LSM 800, Zeiss).

### Human myometrium cell culture

The full-thickness uterus was obtained from women undergoing a scheduled term (≥ 37 weeks of gestation) cesarean section delivery. Patients who lacked signs of infection or known pregnancy complications and were clinically defined as not in labor on the basis of intact membranes, a closed cervix and a quiescent uterus. After delivery of the placenta, samples of myometrium (1cm^3)^ were excised from the upper incisional margin of the lower uterine segment and immediately washed in ice-cold PBS. Myometrium was carefully minced into small pieces of about 1 mm^3^, subsequently washed and incubated with gentle agitation for 40 min, at 37 °C, with collagenase IA (C9891, Sigma-Aldrich) and collagenase XI (C7657, Sigma-Aldrich) each at 0.5 mg/mL in DMEM/F-12 media (A4192001, Gibco) supplemented with BSA (sh30087.02, HyClone) at 1 mg/mL. The dispersed cells were separated from non-digested tissue by filtration through a cell strainer (70 μm, 431751, Corning) and collected by centrifugation of the filtrate at 3000 rmp for 5 minutes at room temperature. The cells were suspended in DMEM/F-12 media supplemented with 10% FBS (10270-106, Gibco) and 1% penicillin/streptomycin/amphoterin B (15240062, Gibco). Cells were then dispersed in 12.5cm^2^ plastic culture flasks (12631, JETBIOFIL) or on glass coverslips into 12-well plates (7516, JETBIOFIL). Culture medium was changed every 48 hours. Cells were maintained at 37 °C in a humidified atmosphere (5 % CO2 in air) until they were semi-confluent (usually 9 days after plating) when the cells were detached using 0.25 % trypsin-EDTA (15050065, Gibco). Experiments were performed using cells between passages 1 and 4. Culture media were collected after incubations and protein concentration of multiple secreted cytokines was analyzed by ELISA.

### Cell interference

To elucidate the involvement of IL-33 in the regulation of uterine contractions during delivery, knockdown experiments were performed in myometrium cells. After starving the cells for 2 hours in serum-free medium, a Lipofectamine 2000 (11668027, Thermo Fisher Scientific) and siRNA IL-33 (Gene Pharma) mixture was added according to the manufacturer’s instructions. After incubation in the incubator for 6 hours, the medium was replaced for a further 48 hours and the efficiency of siRNA transfection was measured by western blotting analysis.

### Medicine loading

Some cells were incubated with lipopolysaccharides (LPS L8880, Solarbio). 10 ug/mL of LPS was added to treat the cells for 5, 10, 15 and 30 minutes as well as 1, 3, 6, 9, 12, 18 and 24h respectively whereas the untreated uterine smooth muscle cells were used as a control group. 4μ8C (80μM, HY-19707, Master of Small Molecules) was used to blocked ER stress and BAPTA-AM (25μg of MAB120503, Abcam), a membrane-permeable calcium ion chelator was used to block the action of cytoplasmic calcium in the cells. These were added to the cells 1 hour before LPS treatment.

### Enzyme-linked immunosorbent assays (ELISA)

Sandwich ELISA were used to determine the cytokine concentrations in culture media following various treatments. ELISA kits for human IL-6 and IL-8 were purchased from MEIMIAN (MM-0049H1 and MM-1558H1, respectively). Culture media were diluted 1:5 for IL-6 and IL-8 determinations using diluent solutions supplied by the manufacturer to ensure the absorbance reading would remain within the range of the standard curve. Absorbance readings were conducted using μQuantTM software (128C-400, BioTek® Instruments) according to the manufacturer’s instructions.

### Statistical analysis

The data are presented as mean percentages of control ± SD. The data from western blots are presented as means ± SEM of the ratios. Statistical analyses were compared using an unpaired two-tailed Student’s t test or a one-way analysis of variance and Bonferroni multiple comparisons test with SPSS software (version 25.0 for Windows, IBM Inc, Chicago, IL, USA) and the figures were generated using Graphpad Prism.

## Data Availability

All data generated or analyzed during this study are included in the manuscript and supporting files.

## Ethics Statement

This study was approved by the Ethics Committee of the First Affiliated Hospital of Nanjing Medical University, China, and complies with the Declaration of Principles of Helsinki.

## Acknowledgement

The authors thank Hong Zhou for advice and technical assistance in early of this work. This study was supported by grants from the National Natural Science Foundation of China (Nos. 81300507).

## Figure legends

**Table 1.**
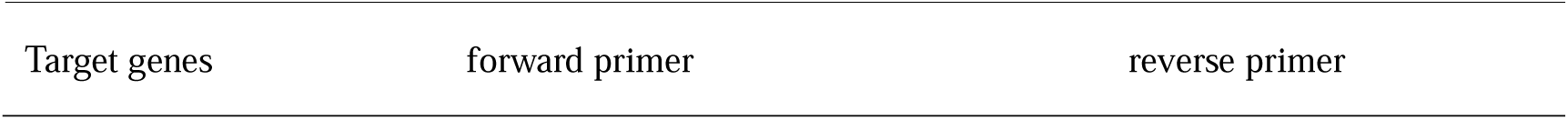
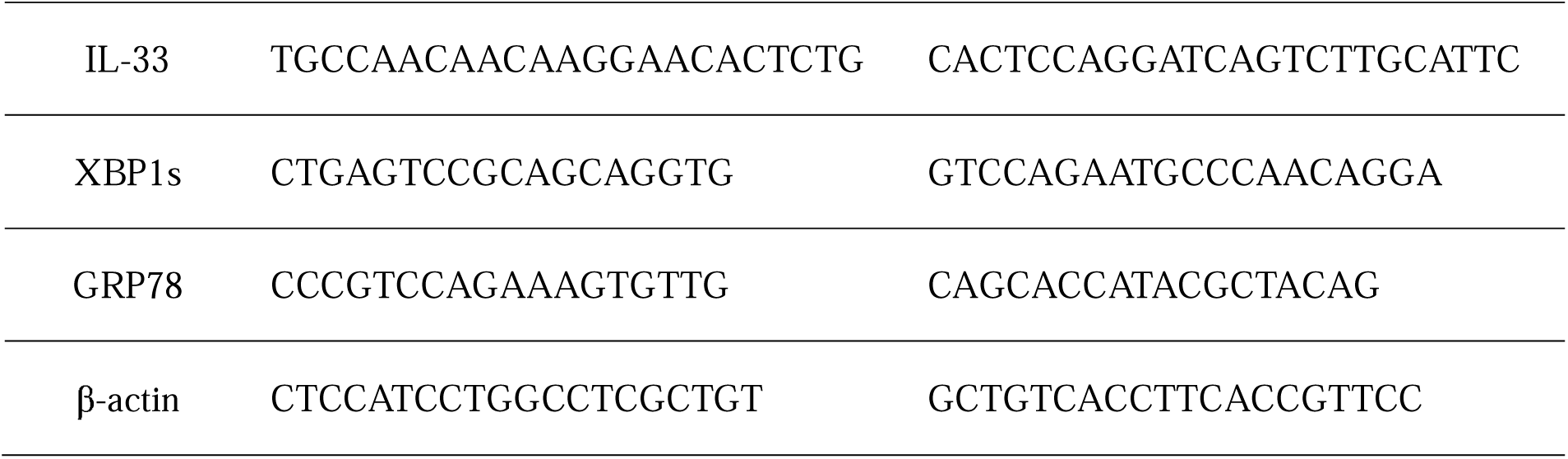
(The following primers are human)

